# Evidence of Unreliable Data and Poor Data Provenance in Clinical Prediction Model Research and Clinical Practice

**DOI:** 10.64898/2026.02.24.26347028

**Authors:** Alexander D Gibson, Nicole M White, Gary S Collins, Adrian G Barnett

## Abstract

Clinical prediction models are often created using large routinely collected datasets. It is essential that prediction models are developed with appropriate data and methods and transparently reported to ensure that decisions are based on reliable predictions. Kaggle is a popular competition and data repository website where users learn and apply analysis skills on a range of datasets. We identified two large, publicly available Kaggle datasets, on stroke and diabetes, that lack clear data provenance, but are widely used in clinical prediction models in peer-reviewed publications. The authenticity of both datasets could not be verified and have no reliable provenance of authenticity and should not be used for informing research or practice. Data provenance assessment using nine TRIPOD+AI items revealed major deficiencies, with minimal details for either dataset including no information on when, where, why or how the data were collected. From these two datasets, we found 125 clinical prediction model studies. Three prediction models had evidence of use in clinical practice, one model was cited in a medical device patent, and the models were cited in 86 review articles. We recommend that journals and data repositories mandate data provenance reporting to safeguard published research. Prediction models based solely on inauthentic or unreliable data sets should never be used to directly inform decisions on patient care.

## Background

Clinical prediction models have become a major area of research with an estimated nearly 250,000 models published in the literature up until 2024 (1). These tools can aid clinicians in patient diagnosis or prognosis that may influence treatment plans and subsequent health outcomes (2). It is essential that clinical prediction models are developed with appropriate data and robust methods to avoid potential misguided clinical decisions and patient harm if poorly developed models are used in practice.

Clinical prediction models must be fully reported to ensure that researchers, clinicians and policy makers can rigorously assess a model for clinical use. The Transparent Reporting of a multivariable prediction model for Individual Prognosis or Diagnosis (TRIPOD) was openly published in 2015 as an expert led checklist of essential items required to assess a model’s clinical utility (3,4). The TRIPOD statement was updated in 2024 to reflect methodological advances, and now also covers models developed using both regression and machine learning approaches and renamed the TRIPOD+AI statement (5). Clinical prediction models that are published without the required minimum level of information can conceivably lead to poor quality outcomes that at best are wasteful and at worst place patients at risk if used in practice. Necessary information includes data provenance, which is the “metadata that confirms the authenticity of that data and enables it to be reused”, including the documentation of where, why, how, when and by whom it was collected (6).

A recent positive trend in health and medical research is the rise of large health and medical datasets, often from routinely collected sources and aided by the growth of automated hospital and patient monitors. These large datasets are widely used to provide insights that improve patient care (7).

However, some large data sets have been used for fast-churn research, where the authors’ goal is to rapidly publish papers rather than improve understanding or health outcomes (8–10). This approach can lead to false discoveries and waste research resources. In this paper, we highlight an emerging issue that combines fast-churn research with poor data provenance and clinical prediction models.

Some publishers have begun to address the problem of fast-churn research using publicly available datasets. Two publishers *PLOS* and *Frontiers* have both updated their policies to ensure the data used by articles meet good standards regarding data quality (11,12). Further, journals including *Scientific Reports* and *Expert Opinion on Drug Policy* have developed individual policies for rejecting fast-churn research (13). These changes to data policies have been spurred by the misuse of the Global Burden of Disease dataset and the National Health and Nutrition Examination Survey, two publicly available datasets which have experienced an explosion of formulaic fast-churn research articles (14). Another example of poor data provenance is the growth of non-verifiable cell lines in the cancer literature, leading to numerous retractions of peer reviewed articles (15).

There are established guidelines, policy documents and checklists that address data provenance integrity issues. The Findable, Accessible, Interoperable and Reusable (FAIR) principles were published in 2016 to improve the management and stewardship of scientific data (16). More recently, the Royal Statistical Society and American Statistical Association launched the “Real World Data Science” project, updating their submission guidelines to include recommendations for transparent data reporting (17). As dataset availability continues to grow, it is essential that data are appropriate for their intended purpose and that key information is reported, including when,where, why and how the data were collected. All secondary uses of data should be pre-registered and analysed appropriately to mitigate against any potential misuse, false discoveries and inappropriate clinical advice.

Online data repositories encourage good research and data practices by facilitating transparency; however, data authenticity is also vital for good research and to the best of our knowledge no data repository or publisher have a mandatory process for reporting data provenance or authenticity.

Kaggle, a popular data repository for machine learning and AI competitions has more than 28 million users and hosts over 550,000 datasets (18). The site encourages skills development, and rewards participants with monetary prizes, badges and recognition. However, Kaggle – like other repositories – allows users to upload datasets without requiring information on data provenance (19). In one recent case, a dataset was uploaded to Kaggle without any ethical consideration for the study participants and was used in multiple research articles that have since been retracted (20).

After inspecting publicly available datasets on Kaggle, we identified serious concerns about the authenticity of several health and medical datasets with suspected poor provenance. We are concerned that medical research is being conducted and published using unreliable data which undermines its relevance for evidence-based medicine and clinical application.

## Methods

### Kaggle datasets

Two publicly available health datasets that could be used to develop clinical prediction models were identified with likely poor data provenance. We arbitrability chose these two datasets from amongst dozens available as they were popular, having been downloaded more than 350,000 times together, and hence were more likely to have been used in research publications. The primary use of each dataset based on their description was predicting health events and hence aligned with our interests in improving clinical prediction model research. One dataset concerned stroke and one concerned diabetes and were accessed and downloaded on 27/08/2025 from Kaggle (https://www.kaggle.com/), links to the Kaggle pages are in Supplementary material 1. The aim of this study is to examine and highlight data provenance issues using two datasets from Kaggle that focus on clinical prediction models.

The two datasets were examined for provenance using the TRIPOD+AI assessment and for authenticity using exploratory analyses of the datasets. We scored each dataset against nine items from the TRIPOD+AI statement that are related to data provenance (Supplementary 1). In addition, we made subjective assessments using all information available on the Kaggle dataset webpages.

We conducted exploratory analyses to look for signs if the data were simulated or fabricated. Patterns between the ID variable and other variables were of interest, as were correlations between variables that are not expected to have correlations, and abnormal distributions of variables (e.g., uniform distributions or erroneous distributions) and duplicate rows.

We did not approach the Kaggle users who uploaded the datasets, but we did examine publicly available discussion posts between all Kaggle users that concerned data provenance. We raised our concerns with Kaggle about the potential harms being caused by the two data sets.

### Study selection

We searched Google Scholar using identifiable terms from the Kaggle web addresses to find research outputs that referenced either dataset. Our complete search terms are in Supplementary material 1. We did not search other bibliometric databases as the search terms did not retrieve relevant outputs (e.g. PubMed). We then screened the full text and references to verify that the Kaggle dataset was used. Only peer-reviewed articles that used the Kaggle datasets in a clinical prediction model development or validation study were included. We excluded conference abstracts, preprints, theses, books, articles without a PDF available, and articles that were not in English. Our search relied on the authors including a link to the Kaggle data and will miss articles that used the data but did not include the Kaggle web site.

One author (AG) screened all eligible articles and assessed the included articles in a random order against the nine TRIPOD+AI checklist items. Another author (AB) conducted a second assessment on a random sample of 10 included articles. Differences in screening outcomes were present for TRIPOD+AI checklist item seven and after discussion AG re-screened checklist item seven for all included articles.

### Clinical Prediction Models

We documented inconsistencies where an item was scored “yes” in the article but “no” for the corresponding the dataset, which would be impossible if the authors relied solely on the available metadata. We checked whether the articles attempted to disclose the origins of the datasets. All articles were reviewed for statements on ethical approval and the Declaration of Helsinki.

Each article was examined for statements that the model has the potential to be used in practice or has been used in practice. We read the full text of the articles to examine if the authors made these recommendations that their models could improve practice, health outcomes or be implemented into clinical practice. We checked if any clinical prediction models had been used in policy by examining Altmetric (https://www.altmetric.com/) and Overton (https://www.overton.io/).

To examine if the problems were concentrated by country, we collected the country affiliation of the first author. To examine trends over time we plotted the volume of research published by time assessed by articles available in OpenAlex (https://openalex.org/).

### Analysis methods

Our study design and analysis methods were pre-registered on the AsPredicted website (pre-registration number: 242613)(21).

In addition to our pre-registered analyses, secondary analyses examined total citation counts per included article and whether they were cited by published reviews, accessed through OpenAlex. We also compiled a tree map of all publishers to assess if the articles using these datasets are being published by specific publishers.

All our data and code are publicly available at https://github.com/alexdgibson/cpm_data_prov (22). All analyses were undertaken in *R* (version 4.5.2) with primary R packages used *tidyverse* (version 2.0.0) and *openalexR* (version 2.0.2).

This study uses publicly available data and does not require a full ethical application and received ethics exempt approval from the Queensland University of Technology on the 28^th^ April 2025, Project ID 9878.

## Results

Searches were carried out in Google Scholar on 15^th^ of August 2025. A total of 653 research outputs were identified of which 125 met the inclusion criteria of being a published clinical prediction model study using either of the two datasets (Supplementary 1). There were 167 conference abstracts, 32 pre-prints, 20 theses, five books and one poster which were excluded as we were interested in published articles. Twelve research outputs were not accessible, and 39 articles did not have a PDF accessible or were not in English. There were 103 articles using the stroke dataset, 20 articles using the diabetes dataset, one article that used both datasets and one article that was retracted using the stroke data. The most recent article was published on 14^th^ August 2025, one day before the search was conducted.

### Dataset provenance

A review of the two Kaggle dataset webpages showed unreliable information pertaining to data provenance. The stroke dataset scored “no” to all nine TRIPOD+AI items in Table 1 (0 out of 9). The diabetes dataset scored “no” to all nine TRIPOD+AI items (0 out of 9).

Information on the Kaggle webpages from the users that uploaded the datasets demonstrated that data provenance was insufficient for use in clinical research. The uploading user of the stroke prediction dataset stated (Supplementary 3):

*“In addition, because the source of the dataset is private, you should only use this dataset for educational purposes, not for research or economical purposes.”*

The uploading user of the diabetes prediction dataset noted:

“*To create the Diabetes Prediction dataset, EHRs were collected from multiple healthcare providers and aggregated into a single dataset. The data was then cleaned and preprocessed to ensure consistency and remove any irrelevant or incomplete information.*”

and

*“I apologize, but due to confidentiality reasons or other restrictions, I’m unable to disclose the specific source of the*

*diabetes prediction dataset.”*

### Exploratory analysis: data irregularities

The stroke prediction dataset contains 5,110 observations from 5,110 unique patient IDs and 12 variables. It has been downloaded from Kaggle more than 275,000 times. Our exploratory analyses identified abnormal patterns with the patient identifier variable and blood glucose variables in the stroke dataset, with a clear shift in the distribution (Figure 2A). Missing data were only present for the body mass index (BMI) variable with relatively few missing values which is unusual for routinely collected health data (23). Only 0.3% of all data points were missing, which is extremely low compared with other health data sets (24). The maximum patient ID was 72,940 while only 5,110 observations were included in the data leaving 93% of potential observations unaccounted for.

**Figure 1.**
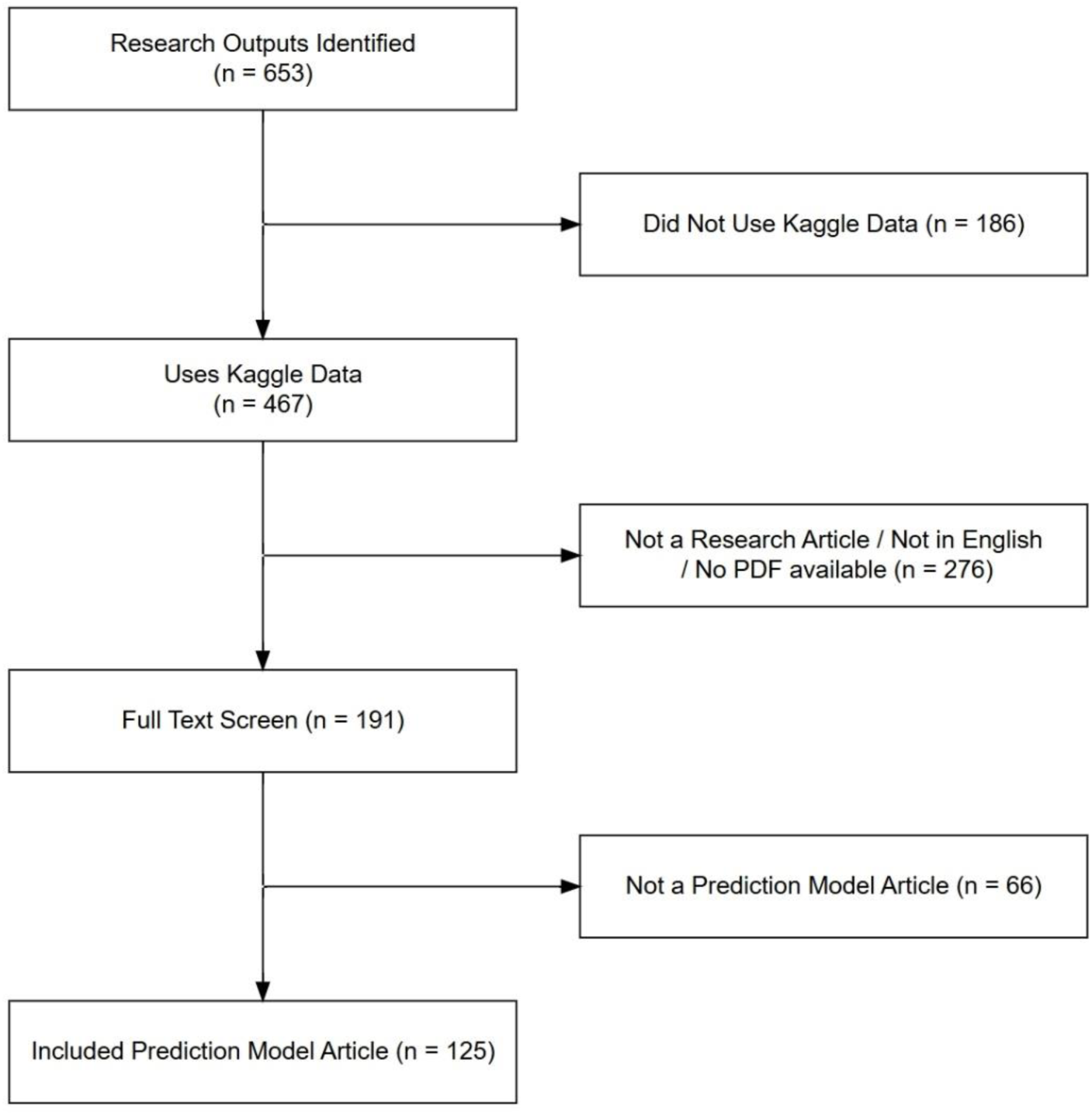
Flow chart of the screening for study inclusion and exclusion.

**Figure 2.**
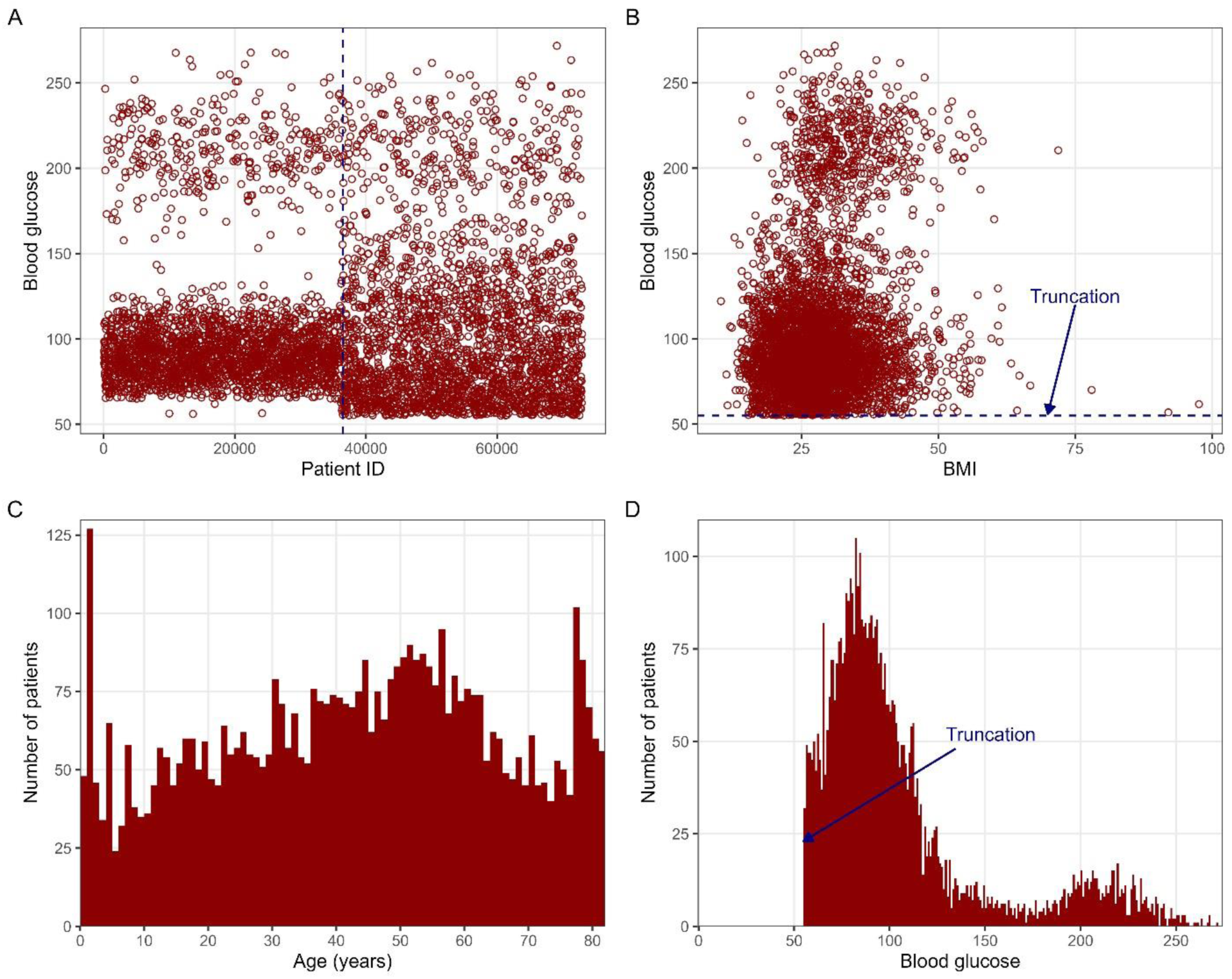
Stroke dataset. “A” Average blood glucose plotted against the patient identifier. “B” Average blood glucose plotted against BMI. “C” Histogram of age with bin widths of 1. “D” Histogram of blood glucose with bin widths of 1. Panel A shows a clear shift in the distribution of blood glucose at half the maximum ID variable. Panels B and D show an unexplained truncation of a blood glucose at 55. Panel C shows an unexplained over-representation of young and old patients, with an unexplained truncation at 82. Note, we have not added the units for BMI or Glucose as these were not in the metadata.

There were abnormal and unexplained for truncations in the blood glucose variable and the maximum age (Figure 2B and 2C).

The diabetes prediction dataset includes exactly 100,000 patients and nine variables. It has been downloaded from Kaggle more than 114,000 times. Visual inspections showed an abnormal and extremely implausible association between BMI and blood glucose (Figure 3A), where a strong association would be expected. There were 18 discrete average blood glucose and HbA1c values for 100,000 patients and shows extreme repetitive and organised patterns indicative of alteration (Figure 3D). This association is not clinically plausible for real data of 100,000 patients and indicates that observations were very likely resampled to create “new” patients. There were highly unusual spikes in the distribution of the patients’ BMIs (anomalously frequent at (27,28]) and ages (anomalously frequent at (1,2] and (79,80]) (Figure 3B and 3C). There were 6,939 (7%) duplicated patient observations which may have been resampled. The blood glucose, HbA1c and BMI variables are biologically implausible and are likely synthetic or fabricated.

**Figure 3.**
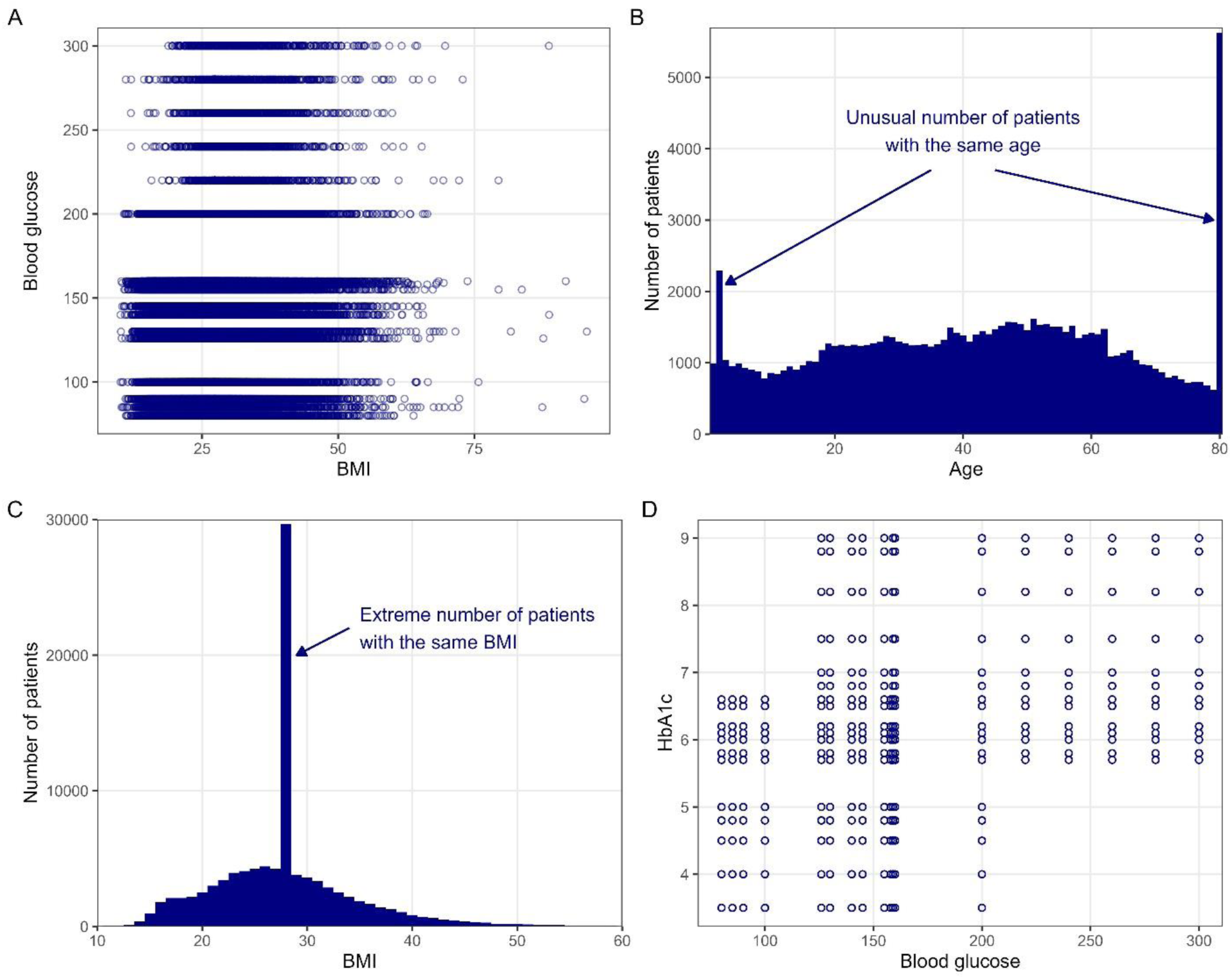
Diabetes dataset. “**A**” Blood Glucose plotted against BMI. “**B**” Histogram of age. “C” Histogram of BMI in bin widths of 1 unit. “D” HbA1c level plotted against blood glucose level. . There were just 18 discrete results for blood glucose and HbA1c which is not plausible in real health data and shows erroneous patterns. Close to 30,000 patients had a BMI of (27,28] units with the next largest frequency at just over 4,400 patients. Note, we have not added the units for BMI, HbA1c or Glucose as these were not in the metadata.

**Figure 4.**
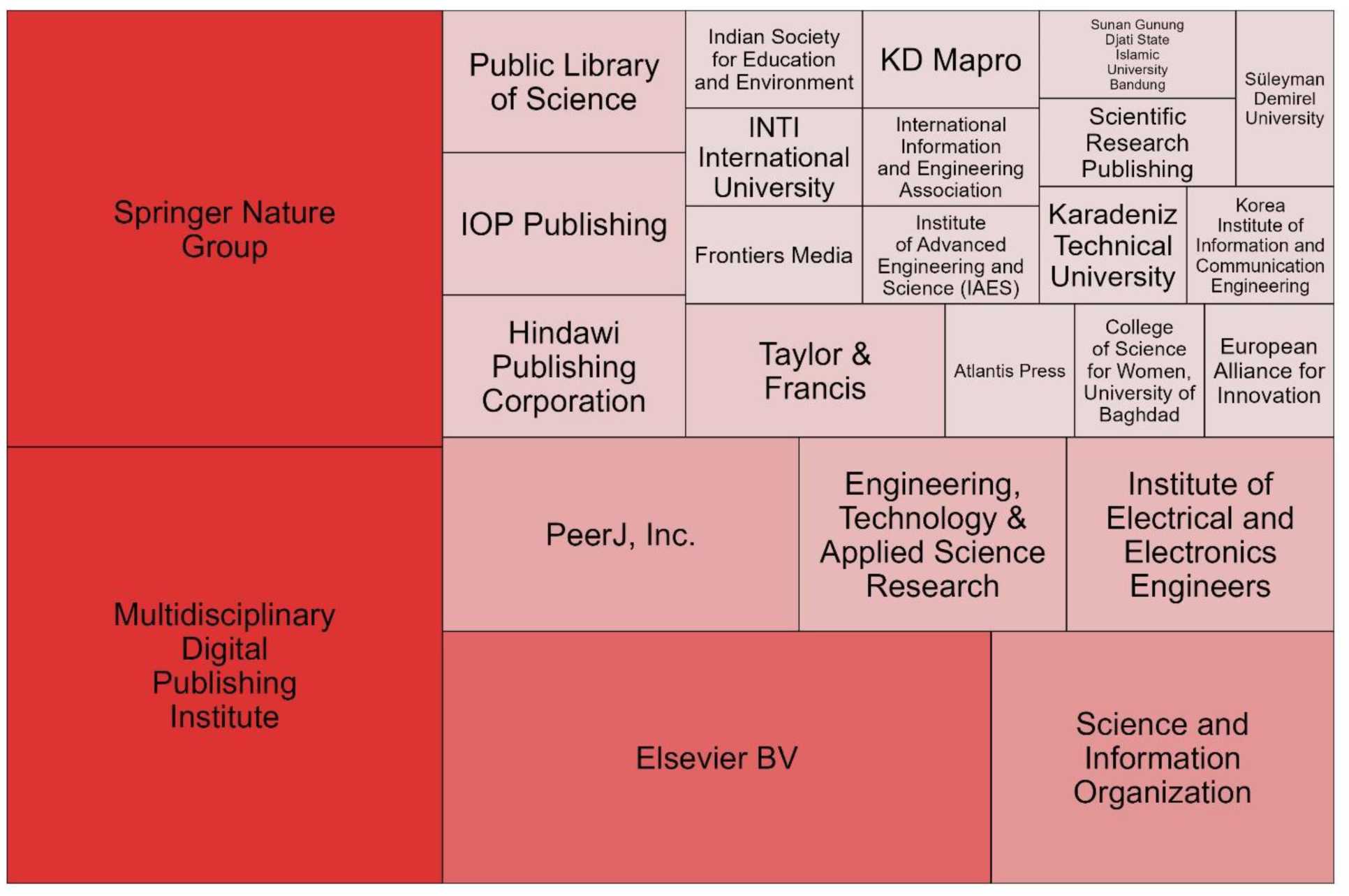
Tree map of the publishers that published one or more clinical prediction model using one of the two Kaggle datasets as assessed by OpenAlex. Of the 125 articles, 108 were available in OpenAlex and 67 of those had publisher data available in OpenAlex.

### Research Articles: Descriptive results

Authors from 32 different countries developed clinical prediction models using either of the two datasets, assessed by the first author country affiliation. The top 10 most prominent countries are shown in Table 2.

**Table 2.** The top 10 countries that have published an article using the diabetes or stroke prediction datasets by the first author’s country affiliation.

| Country | Number of Clinical Prediction Model Articles |
| --- | --- |
| India | 31 |
| Indonesia | 15 |
| China | 12 |
| Saudi Arabia | 10 |
| Bangladesh | 8 |
| Iraq | 7 |
| Turkey | 5 |
| United Kingdom | 4 |
| United States of America | 4 |
| Malaysia | 3 |

Only 2% of articles (n=3) had a statement that ethical approval was approved or that ethical procedures were followed which is a reporting item in TRIPOD+AI (Table 3). Only 7% (n=9) had a statement that ethics was not required. Most articles (90%) had no ethical statement at all.

**Table 3.** Number of research articles that provided a statement of ethics or ethical considerations, statement that ethics not required, no statement as well as mentioning the Declaration of Helsinki.

| **Data set used** | **Statement of Ethics** | **Statement Ethics Not Required** | **No Ethics Statement** | **Mention of the Declaration of Helsinki** | **Total** |
| --- | --- | --- | --- | --- | --- |
| Stroke | 2 | 7 | 95 | 0 | 104 |
| Diabetes | 1 | 2 | 17 | 0 | 20 |
| Both | 0 | 0 | 1 | 0 | 1 |
| Total | 3 | 9 | 113 | 0 | 125 |

Adherence to the nine TRIPOD+AI items on data provenance was poor across the 125 included studies (see Table 4). There were only 76 (7%) items reported from all articles out of a total 1,125 items.

**Table 4.** Results of adherence to the nine TRIPOD+AI items on data provenance using the information available for the two Kaggle data sets and the information provided by published articles.

| TRIPOD+AI<br>ITEM | 5a | 5b | 6a | 7 | 8a | 9a | 9b | 10 | 11 |  |
| --- | --- | --- | --- | --- | --- | --- | --- | --- | --- | --- |
| Assessment of Kaggle data sets |  |  |  |  |  |  |  |  |  | Total<br>TRIPOD+<br>AI item<br>count |
| Stroke data | 0 | 0 | 0 | 0 | 0 | 0 | 0 | 0 | 0 | 0/9 |
| Diabetes data | 0 | 0 | 0 | 0 | 0 | 0 | 0 | 0 | 0 | 0/9 |
| Assessment of published articles |  |  |  |  |  |  |  |  |  | n / N |
| Stroke Articles<br>(n=104) | 5 | 0 | 2 | 1 | 0 | 1 | 0 | 0 | 63 | 72/936 |
| Diabetes<br>Articles (n=20) | 0 | 0 | 0 | 0 | 0 | 1 | 0 | 0 | 2 | 3/180 |
| Both (n=1) | 0 | 0 | 0 | 0 | 0 | 0 | 0 | 0 | 1 | 1/9 |
| Item Totals<br>(n=125) | 5 | 0 | 2 | 1 | 0 | 2 | 0 | 0 | 65 | 76/1,125 |

**Table 5.** Recommended minimum mandatory data provenance reporting list for data repositories using health and medical research.

| Question | Answer |
| --- | --- |
| What kind of data? | Real Data / Simulated Data / Synthetic Data / Comment Other. |
| If data are synthetic or simulated, how so? | Provide code that generated the data. |
| Has any of the data been processed?<br>If yes, which variables and how. | Yes / No<br>If yes, list which data are processed and how. |
| Were any data removed? Including individual observations, whole variables or patients. | Yes / No/ Unsure<br>If yes, list which data were removed and why. |
| Who collected the data? | List the institution(s) that collected the data as well as the number of individuals. |
| What data were collected? | List all the variables, including the machines, techniques, measurements, units, software or processes by which the data were collected. |
| When was the data collected? | Start and end dates of data collection, as well as any interruptions to data collection. |
| Where were the data collected? | Geographical location(s) of the data collection. |

Five articles using the stroke dataset made a broad attempt at describing the source of the data, item 5a of the TRIPOD+AI checklist (Supplementary 2). Two of these articles stated the data came from “Bangladesh clinics” [Stroke Article 29 and 72; see supplement 2] with no additional information. One article stated the data were from “prestigious healthcare organizations and institutions like AIMS and WHO” [Stroke Article 39]. One article stated the data came from “clinical volunteers” [Stroke Article 47]. One article stated the data was from “McKinsey & Company EHR [electronic health record]” [Stroke Article 60]. No diabetes prediction articles included information on where the data originated from.

Two articles provided a description of location of study setting (item 6a), these were the two articles which identified that data came from Bangladesh clinics. Only one article using the stroke dataset scored “yes” to item 7 for pre-processing and quality checking as they mentioned the data do not come from a peer-reviewed clinical study or government registry and there is a lack of documentation, concluding it is potentially synthetic or anonymised [Stroke Article 36]. Two articles described the choice of initial predictors and their pre-selection (item 9a), whilst 65 articles mentioned missing data (item 11) as they provided some information on handling of missing data.

As of 21^th^ April, 2026, 16 articles that used the diabetes dataset (80%) were indexed on OpenAlex. Ten of the diabetes articles had one or more citations and received a total of 84 citations, including being cited in three review articles. Ninety-one of the 104 articles using the stroke dataset (88%) were indexed in OpenAlex. Seventy-six of these articles had one or more citations and received a total of 1,695 citations, including being cited in 83 review articles. The one article that used both datasets had been cited two times and not cited in any review articles.

### Clinical Recommendations

Practical recommendations were reported in 70 of 104 (67%) prediction model studies using the stroke dataset and 16 of 20 (80%) studies using the diabetes dataset. The one article using both datasets made no practical recommendations.

Examples of practical recommendations from articles were (article identifiers described in supplementary 2):

· “Collectively, these techniques contribute to robust, reliable, and more practical machine learning models suitable for real-world applications. The healthcare system may see less strain and better patient outcomes as a result of this in the long run.” [Stroke Article 44]
· “Therefore, healthcare providers can use these classifiers that are the most suited for predicting stroke based on the medical history of a patient in the real world.” [Stroke Article 86]
· “The results of this study will not only help individuals receive the necessary medical interventions in a timely manner to reduce the incidence of diabetes mellitus and its

complications, but will also have a significant impact on the health and quality of life of the entire middle-aged and elderly population.” [Diabetes Article 4]

There was potential evidence that three of the clinical prediction models have been used or will be used in clinical practice on patients as mentioned in the articles. Examples of potential use in clinical practice from articles (Supplementary 2 [Stroke Articles 17, 56, 69]):

· “Therefore, the quality of medical treatment for stroke patients at RSUD Banyumas is expected to improve, and the mortality rate caused by stroke will decrease.” [Stroke Article 17]
· “As a use case, one of these models has been successfully tested in diagnosing stroke events” and “We conducted several experiments with five patients of different ages, in order to evaluate the accuracy of the results obtained and the evolution of the prediction of each patient in developing a stroke event based on the demographic values and laboratory tests performed.”[Stroke Article 69]
· “Additionally, the study’s validation process relied exclusively on internal validation through training and test splits. To ensure the model’s robustness and generalizability in real-world clinical practice, external validation is imperative. Ongoing efforts include deploying the prediction model in a local heart clinic to validate its performance in clinical settings.” [Stroke Article 56]

There were an additional four articles (Supplementary 2 [Stroke Articles 16, 38, 49, 90]) that had a statement that the models may have been used in practice but lacked additional evidence in the article to support these models were genuinely used.

Eleven clinical prediction models contained computer-based or phone-based tools with graphical user interfaces (Supplementary 2 [Stroke Articles 21, 26, 31, 38, 55, 65, 69, 81, 91, 101 and Diabetes Article 19]); two of these are publicly available and accessible [Stroke Articles 38 and 55].

None of the clinical prediction model studies were found to be referenced in policy documents, as assessed by Altmetric and Overton. One stroke article was referenced in a medical device patent from the California Institute of Technology and University of Southern California for the physiological metrics for determining stroke risk (Supplementary 2 [Stroke Article 104]).

There were 108 (86%) out of the 125 articles accessible for publication dates on OpenAlex with seven articles not having DOIs and 10 articles with DOIs not being indexed in OpenAlex. Since 2021 there has been a continuously increasing volume of publications using the datasets (Figure 5).

**Figure 5.**
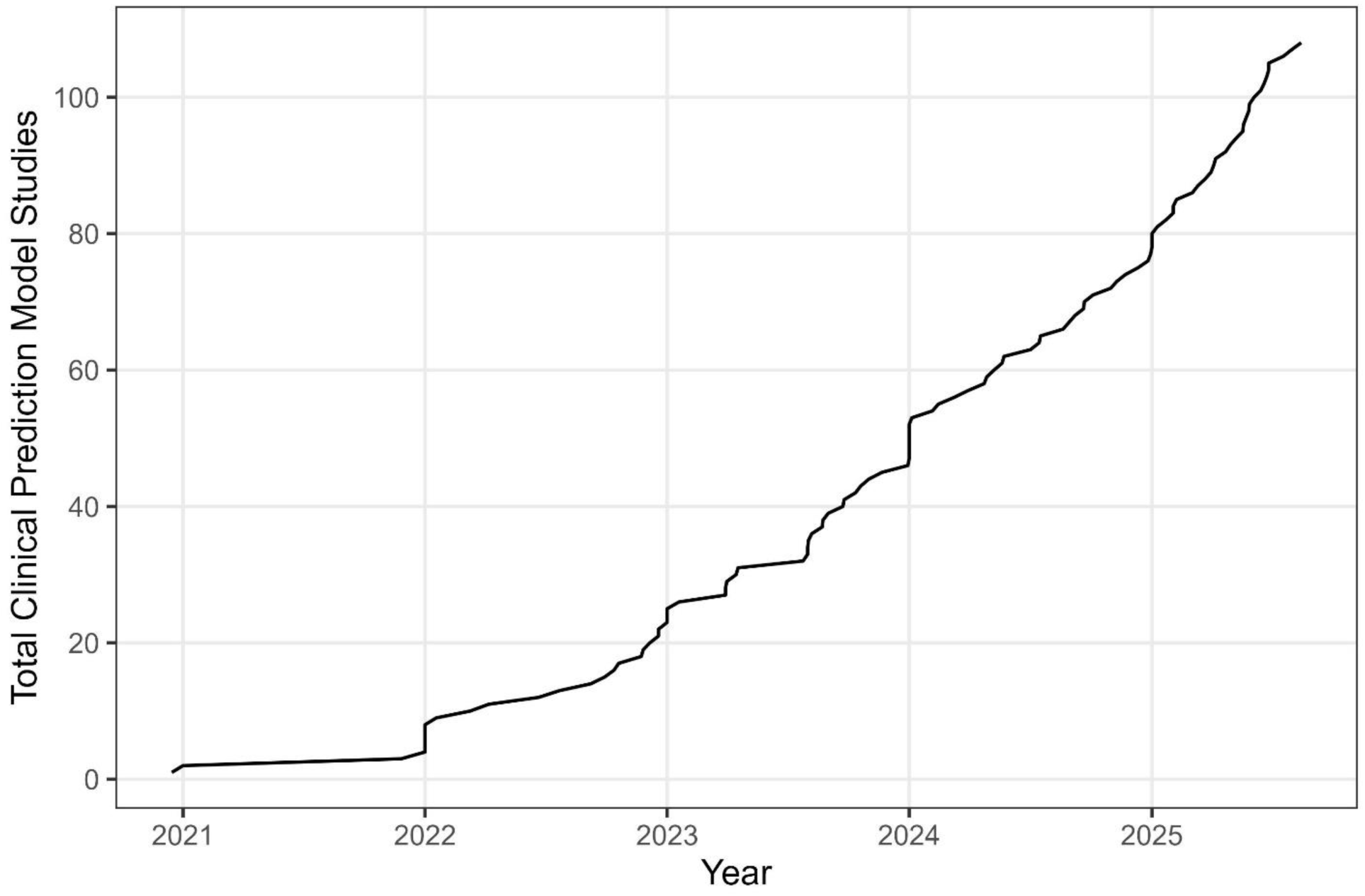
Cumulative number of publications using either of the two Kaggle datasets over time assessed by OpenAlex publication date. There were 108 of the 125 articles available in OpenAlex with a publication date available.

## Discussion

Our research has found that clinical prediction models are being developed and validated using unreliable publicly available data with poor provenance. Worryingly, the results are being used to inform further research via their uptake in reviews. The articles indexed in OpenAlex have received a total of 1,781 citations, including citations in 86 review articles and a medical device patent for stroke risk. These numbers show that the articles are informing other research and potentially misdirecting future work with no signs of slowing down (Figure 5). Both datasets examined had extremely poor data provenance as assessed by nine TRIPOD+AI items. Poor data provenance could be attributed to the lack of mandatory reporting in data repositories. Additionally, there were statements from both users who uploaded the datasets stating that the source of data cannot be disclosed. Both datasets show strong signs that they are unreliable with no data provenance and unexplained data irregularities. Even with such poor provenance, 125 clinical prediction model studies were published with practical recommendations being made in 67% of stroke articles and 80% of diabetes prediction articles. This result is similar to previous research on spin practices in prediction models which found that 86% of articles recommended use in clinical practice; however, those models were presumed to be based on real data(25).

Of great concern is evidence that three of these clinical prediction models have potentially been used in clinical practice. If clinical decisions are being guided by prediction models that are developed with data that is not fit for purpose it can place patients at increased risk of harm as poor predictions will lead to patients being denied necessary treatments or receiving unnecessary treatments. Eleven articles developed digital prediction tools with a graphical user interface for mobile or web applications and two of these are currently publicly accessible [Stroke Articles 38 and 55] and these may be being used in practice.

Ideally, all the online tools based on the Kaggle datasets would be immediately removed until the data provenance can be assured. All the clinical prediction model articles should have an expression of concern placed on them until verification of data can be made and any additional articles using these datasets should not be accepted for publication. If these datasets cannot be verified, research using these articles with the assumption the data is real should be retracted. Since we flagged the articles on PubPeer (https://pubpeer.com/) in March 2026, three articles in Scientific Reports have been retracted for the unreliable use of the stroke data and two more have an expression of concern placed on them. One article was already retracted before we began our analysis for, among other things, concerns about authorship, potential “tortured phrases” (26) and citing retracted work (27). Taking action to remove the datasets from Kaggle is important as more articles are likely in the process of being published (Figure 5). The most recent article was published one day before our search was completed.

Data provenance information was inconsistent between articles that used the datasets and the information provided by the data repository. Five stroke articles identified four unique sources for the data which cannot be verified and at a minimum three must be wrong. Two stroke articles described the clinical setting; however, this information was not provided in the data repository.

Two articles included additional information that was not in the Kaggle repository, as one stroke article described quality checking, and one article from the stroke and diabetes datasets identified a pre-selection of predictors. It remains unclear how some authors and articles were able to include such detailed additional information when this was not provided in the data repository. The additional information may have been fabricated to pass peer review, adding illusory provenance and false authenticity to the data.

Only one article scored “yes” to TRIPOD+AI item 7 for checking data quality, identifying that the data are not from a valid source. Nonetheless, even though the article identified the data was of poor provenance, the authors still made recommendations for the potential practical implications of the model.

It was clear from the statements on the two Kaggle dataset web pages that these datasets have no information available to verify the data with other Kaggle users questioning authenticity and validity (Supplementary 3). Both dataset descriptions mention that the source cannot be disclosed and for the stroke dataset it is explicitly stated not to be used in research. These issues raise serious concerns as to the use of these models in research and practice as the underlying data from available information is not reliable. Exploratory analyses of the diabetes dataset indicated patterns that were consistent with being simulated or fabricated, and both datasets had a remarkable lack of missing data. We cannot however conclude with certainty that either of the datasets are entirely simulated or fabricated. As experienced users of health data, we have little confidence that these two datasets are suitable for research given the unexplained lack of missing data, duplicated data rows, incongruitous associations between variables, erroneous truncations and spikes in the data, and complete lack of data provenance.

A striking example of the poor data provenance is that there was no information reported for either the stroke dataset or diabetes dataset on the type of disease. The stroke datasets did not report if the strokes were ischemic, haemorrhagic or both, but haemorrhagic and ischemic strokes both follow different in pathological aetiology and inform different treatment plans. Similarly, Type-1 and Type-2 diabetes both have physiological differences that require unique treatment plans, but this distinction was not made in the diabetes dataset. Further, no information was provided for the units of measurement in either dataset, which is crucial information if a model were to be used in practice.

Only three of the 125 prediction model articles had a statement that ethics was approved for their studies. Ethical approval is essential for research with human participants as defined in the Declaration of Helsinki and section 23 on Research Ethics Committees and must be upheld as fundamental principles for protection of research participants (28). Studies should include reporting of important information, including if ethical approval was required or not. As the study data used in these prediction models is publicly available, ethical approval is likely not needed but only 7% of articles reported that ethical approval was not needed. Still, if these data are real, there is no information pertaining to its collection and it may have been collected or shared without adequate ethical approvals.

We found published articles from 32 countries assessed by the first author’s affiliation, indicating that this is a multi-country issue. Given the increasing growth of publications using these two datasets (Figure 5), it is essential that data provenance issues are addressed by Kaggle. Kaggle’s Privacy Statement states “We use information to help improve the safety and reliability of our Services. This includes detecting, preventing, and responding to fraud, abuse, security risks, and technical issues that could harm Kaggle, our users, or the public.” (29). We have shared our concerns with Kaggle and they have responded that they are working on ways to help inform users of provenance when deciding on source materials. With the increasing use of AI in health and medical research and large datasets being made publicly accessible, it is crucial that data provenance is detailed to prevent unreliable clinical decisions and wasted resources like the ones described in this article. Data provenance is an essential characteristic in assessing clinical prediction models as well as the broader health and medical research area.

We did not approach any of the authors, so we cannot know why so many researchers were willing to publish studies using unreliable data. However, we speculate the authors’ desire to publish a peer reviewed paper has over-ridden concerns about the data. Authors responses on PubPeer indicate that they were willing to accept the datasets as real because they had been widely used in the machine learning community (30,31). Whilst there are many blogs and videos on the technical aspects of machine learning techniques, researchers would be wise to also learn about data provenance and basic sense checks of data quality.

We appreciate the issues we highlight in this article would not have been detectable if it were not for open data sharing practices. It is also important to note that most data sets can be useful for developing statistical and machine learning skills, and can provide useful insights when the data are well described (18,32). These data and code sharing practices are essential for transparency and reproducibility in health and medical research, and we encourage all journals to include mandatory data availability as described in TRIPOD+AI guidelines for clinical prediction models. It was only through such practices these erroneous and abnormal patterns were able to be seen in the datasets.

While this research has highlighted the use of two datasets in the literature, we believe that poor data provenance is likely a wider issue. Our study was limited to two datasets from Kaggle as examples of unreliable data and poor provenance and remains unclear how widespread this issue is within Kaggle datasets and other repositories. There were 276 research outputs identified in Google Scholar that used these datasets but were not included in our assessments as they were not published articles using a clinical prediction model (e.g., book chapters, conference abstracts, theses). Further, there are more doubtful datasets on Kaggle that we did not examine due to time constraints and feasibility. We only searched for articles in Google Scholar referencing the Kaggle web addresses, meaning our sample may be an underrepresentation of use, having missed other research using these two datasets. We are concerned that more datasets with no or very poor data provenance, datasets that are inauthentic, unreliable or at worst, simulated or fabricated, are being used to publish clinical prediction model studies and subsequently being used in practice.

## Recommendations

Given the growing issues identified previously with the NHANES data (14), non-verifiable cell lines (15), image manipulations (33), fraudulent data (34) and now unreliable data, we outline practical recommendations for journals and publishers, data repositories, researchers and clinicians to promote and conduct high quality research.

### Recommendations for Journals and Publishers

Research integrity issues have been growing in the medical literature with some publishers and journals introducing policies and guidelines to prevent inappropriate research from being published and to improve the quality of published research. There are now nearly 700 reporting guidelines with the Enhancing the QUAlity and Transparancy Of health Research (EQUATOR Network). For example established guidelines include the CONsolidated Standards Of Reporting Trials (CONSORT) statement (35) for clinical trials, TRIPOD+AI (5) for clinical prediction models and the Strengthening the Reporting of Observational Studies in Epidemiology (STROBE) statement for observational studies (36).

Our recommendations are similar to previous recommendations from the BMJ and guideline recommendations from the STANDING Together consensus (14,37–39). We also note the recent BEAMRAD tool which was designed to increase the completeness of medical dataset documentation (40). Our recommendations aim to improve research practices in reporting of data provenance.

1. Information on data provenance should be mandatory for all data availability statements regardless of if the data are made publicly available or not, including:

a. What was the primary reason for data collection (e.g. pilot study, randomised control trial, secondary data, retrospective, prospective)
b. Who collected the data (e.g. company, institution, hospital)
c. What data were collected with a clear description of measurements, definitions and units (e.g., a data dictionary)
d. When the data were collected, including start and end dates
e. Where were the data collected, including country, city/region, setting (hospital, outpatient, community, laboratory), and whether single of multiple sites
f. Who funded the data collection
2. Openly available unprocessed data should be mandatory when there are no confidentiality issues preventing data from being made accessible. Accessible data helps researchers contribute to good research practices, allowing the identification of problematic issues as well as verification and reproducibility of results (39).
3. Journals should ensure a data provenance checklist is provided at the time of submission so issues can be identified quickly for early rejection if the data appear to be unreliable. For clinical prediction model studies this can be a TRIPOD+AI checklist.
4. Articles using previously identified synthetic, fabricated or unreliable datasets could automatically be rejected unless the title and abstract state that the data are synthetic, fabricated or lack provenance and the authors include strong justification why the dataset was used.

### Recommendations for Data Repositories

As data are not stored with journals and publishers but instead stored predominately on repositories like, GitHub, Zenodo, or The Open Science Framework, these repositories should add mandatory provenance reporting. We make additional recommendations for repositories to enhance mandatory provenance information for all health and medical research. The goal is to prevent problematic datasets from being used in research and practice.

### Recommendations for Researchers and Clinicians

Both researchers and clinicians need to be aware of data provenance when developing, validating and implementing clinical prediction models. Currently, researchers and clinicians should not assume published research is without errors as poor quality research can pass peer review (41). Prioritising research that is pre-registered, pre-printed and that follows good research practices will help build confidence that the results and clinical implications of a study will be valid and reliable (42). Similar to a data provenance statements, researchers and clinicians should continue to follow best practices for clinical prediction model research and are advised to read the TRIPOD+AI statement (3,5) and PROBAST+AI statement (43).

## Conclusion

This research has identified a growing concern affecting clinical prediction model research with the use of unreliable data from a data repository website. It is essential that clinical prediction models and clinical decisions are made with reliable data and appropriate methods to avoid unreliable health decisions for patients. We have provided recommendations for journals, publishers, data repositories as well as researchers and clinicians to improve research and clinical practice.

### List of Abbreviations

**TRIPOD+AI** is the Transparent Reporting of a multivariable prediction model for Individual Prognosis Or Diagnosis + Artificial Intelligence

**PROBAST+AI** is the Prediction model Risk Of Bias ASsessment Tool + Artificial Intelligence

**EQUATOR** is the Enhancing the QUAlity and Transparency Of health Research

**CONSORT** is the Consolidated Standards of Reporting Trials

**STROBE** is the STrengthening the Reporting of OBservational studies in Epidemiology

**STANDING** is the Standards for Data Diversity, Inclusivity, and Generalisability

**EHR** is Electronic Health Record

**FAIR** is Findable, Accessible, Interoperable and Reusable

## Declarations

### Ethics approval and consent to participate

This study uses publicly available data and does not require a full ethical application and received ethics exempt approval from the Queensland University of Technology on the 28^th^ April 2025, Project ID 9878.

### Consent for publication

Not applicable.

### Availability of data and materials

The datasets generated and/or analysed during the current study are available in the GitHub repository, https://zenodo.org/doi/10.5281/zenodo.18764114

### Competing interests

The authors declare that they have no competing interests.

## Funding

This research was supported by an Australian Government Research Training Program Scholarship https://doi.org/10.82133/C42F-K220 for author ADG. GSC is funded by an MRC-NIHR Better Methods Better Research grant (MR/Z503873/1). GSC is funded by the National Institute for Health and Care Research (NIHR) Birmingham Biomedical Research Centre at the University Hospitals Birmingham NHS Foundation Trust and the University of Birmingham. GSC is a National Institute for Health and Care Research (NIHR) Senior Investigator. The views expressed are those of the authors and not necessarily those of the NHS, the NIHR or the Department of Health and Social Care.

## Authors contributions

AG conceptualised the study. AG and AB collected and analysed the data. All authors helped developed the protocol, analysis plan and final manuscript. All authors have read and approved the final manuscript.

## Data Availability

All our data and code are publicly available at https://github.com/alexdgibson/cpm_data_prov.

## Acknowledgements

We acknowledge Paper-Wizard for providing helpful feedback on a draft of this paper.

## Notes

### Competing Interest Statement

The authors have declared no competing interest.

### Funding Statement

This research was supported by an Australian Government Research Training Program Scholarship doi.org/10.82133/C42F-K220 for author ADG. GSC is funded by an MRC-NIHR Better Methods Better Research grant (MR/Z503873/1). GSC is funded by the National Institute for Health and Care Research (NIHR) Birmingham Biomedical Research Centre at the University Hospitals Birmingham NHS Foundation Trust and the University of Birmingham. GSC is a National Institute for Health and Care Research (NIHR) Senior Investigator. The views expressed are those of the authors and not necessarily those of the NHS, the NIHR or the Department of Health and Social Care.

### Author Declarations

The data for this study was publicly and openly available on www.kaggle.com which were accessed and downloaded on 27/08/2025. The first dataset is: https://www.kaggle.com/datasets/fedesoriano/stroke-prediction-dataset The second dataset is: https://www.kaggle.com/datasets/iammustafatz/diabetes-prediction-dataset

### Summary of Updates

This version of the manuscript has been revised with updated information, analyses and discussion.

