## Supplementary 2 for "Evidence of Unreliable Data and Poor Data Provenance in Clinical Prediction Model Research and Clinical Practice"

List of all the included clinical prediction models articles for the research project using the stroke dataset.

| Reference Title | Article Title | DOI / Web Address |
| --- | --- | --- |
| Stroke Article 1 | Predictive Modeling Of Stroke Occurrence Using Python For Improved Risk Assessment | 10.5937/jpmnt12-50921 |
| Stroke Article 2 | A-Tuning Ensemble Machine Learning Technique for Cerebral Stroke Prediction | 10.3390/app13085047 |
| Stroke Article 3 | Enhancing accuracy in brain stroke detection: Multi-layer perceptron with Adadelta, RMSProp and AdaMax optimizers | 10.3389/fbioe.2023.1257591 |
| Stroke Article 4 | Early Prediction of Stroke Based on Deep and Machine Learning by Applying Medical Imaging and Tabular Data | 10.18280/mmep.111228 |
| Stroke Article 5 | Heart disease severity level identification system on Hyperledger consortium network | 10.7717/peerj-cs.1626 |
| Stroke Article 6 | A comprehensive explainable AI approach for enhancing transparency and interpretability in stroke prediction | 10.1038/s41598-025-11263-9 |
| Stroke Article 7 | An Intuitive Approach on Transfer Learning with an IBF+IHP Model for Stroke Classification and Prediction | 10.48084/etasr.9031 |
| Stroke Article 8 | Stroke Prediction Using Machine Learning Method with Extreme Gradient Boosting Algorithm | 10.30812/matrik.v21i3.1666 |
| Stroke Article 9 | Crunch Mode: Make Early Predictions about Risk of Stroke Using Machine Learning | 10.56977/jicce.2025.23.1.17 |
| Stroke Article 10 | Predicting Stroke Risk with Machine Learning and Hyperparameter Optimization | 10.31466/kfbd.1538305 |
| Stroke Article 11 | Comparative analysis of resampling algorithms in the prediction of stroke diseases | 10.56919/usci.2123.011 |
| Stroke Article 12 | Early Prediction of Stroke Risk Using Machine Learning Approaches and Imbalanced Data | 10.56286/1vf19469 |
| Stroke Article 13 | An Optimization Precise Model of Stroke Data to Improve Stroke Prediction | 10.3390/a16090417 |
| Stroke Article 14 | The Early Warning Signs of a Stroke: An Approach Using Machine Learning Predictions | 10.4236/jcc.2024.126005 |
| Stroke Article 15 | Performance of Machine Learning Algorithms for Heart Disease Prediction: Logistic Regressions Regularized by Elastic Net, SVM, Random Forests, and Neural Networks | https://rrpress.utsa.edu/items/3affdf66-8d85-494c-888e-6fb8e6f181a5 |
| Stroke Article 16 | Predictive Modelling of Stroke Occurrence among Patients using Machine Learning | 10.61453/intij.202355 |
| Stroke Article 17 | Enhancing stroke prediction models: A mixing of data augmentation and transfer learning for small-scale dataset in machine learning | 10.1016/j.cmpbup.2025.100198 |
| Stroke Article 18 | Hybrid Features Binary Classification of Imbalance Stroke Patients Using Different Machine Learning Algorithms | 10.46300/91011.2022.16.20 |
| Stroke Article 19 | Privacy-enhanced heart stroke detection using Federated Learning and Homomorphic Encryption | 10.1016/j.smhl.2025.100594 |
| Stroke Article 20 | Detection of Stroke (Cerebrovascular Accident) Using Machine Learning Methods | 10.17798/bitlisfen.1539189 |
| Stroke Article 21 | Stroke Prediction Using Machine Learning | https://hspublishing.org/JRECS/article/view/372 |
| Stroke Article 22 | An Analysis Of Brain Stroke Prediction Using Machine Learning | 10.61841/turcomat.v11i3.14508 |
| Stroke Article 23 | Design and Development of Modified Ensemble Learning with Weighted RBM Features for Enhanced Multi-disease Prediction Model | 10.1007/s00354-022-00190-2 |
| Stroke Article 24 | Stroke Prediction Using the Trust Evaluation with Data Leakage Avoiding | 10.1088/1742-6596/2560/1/012051 |
| Stroke Article 25 | A Novel Explainable Attention-Based Meta-Learning Framework for Imbalanced Brain Stroke Prediction | 10.3390/s25061739 |
| Stroke Article 26 | Unlocking stroke prediction: Harnessing projection-based statistical feature extraction with ML algorithms | 10.1016/j.heliyon.2024.e27411 |
| Stroke Article 27 | Logistic Regression for Stroke Prediction: An Evaluation of its Accuracy and Validity | 10.54097/hset.v39i.6712 |
| Stroke Article 28 | Optimizing Accuracy of Stroke Prediction Using Logistic Regression | 10.37802/joti.v4i2.278 |
| Stroke Article 29 | Enhanced Stroke Risk Prediction: A Fusion of Machine Learning Models for Improved Healthcare Strategies | 10.1007/s42979-024-03389-w |
| Stroke Article 30 | Navigating Heart Stroke Terrain: A Cutting-Edge Feed-Forward Neural Network Expedition | 10.47738/jads.v6i3.763 |
| Stroke Article 31 | Stroke prediction based on improved machine learning algorithm | 10.1117/12.2659156 |
| Stroke Article 32 | Stroke risk prediction using multiple machine learning algorithms | 10.1117/12.2673750 |
| Stroke Article 33 | Predicting stroke risk: An effective stroke prediction model based on neural networks | 10.1016/j.jnrt.2024.100156 |
| Stroke Article 34 | A Hybrid Machine Learning Approach to Predict the Risk of Having Stroke | 10.1145/3542954.3543020 |
| Stroke Article 35 | Harnessing the hybrid machine learning methods for stroke risk classification | 10.1080/10255842.2025.2501636 |
| Stroke Article 36 | Comparative Evaluation of Machine Learning Models for Stroke Prediction in Clinical Settings | 10.37256/ccds.6220256976 |
| Stroke Article 37 | Fusing Attention and Convolution: A Hybrid Model for Brain Stroke Prediction | 10.4108/eetsis.7022 |
| Stroke Article 38 | A Web-Based Interface That Leverages Machine Learning to Assess an Individual’s Vulnerability to Brain Stroke | 10.1109/ACCESS.2025.3566093 |
| Stroke Article 39 | Brain stroke prediction model based on boosting and stacking ensemble approach | 10.1007/s41870-023-01418-0 |
| Stroke Article 40 | Stroke Disease Detection and Prediction Using Robust Learning Approaches | 10.1155/2021/7633381 |
| Stroke Article 41 | Computational Health Care Analysis Using Hadoop – Stroke Prediction | https://www.researchgate.net/profile/Marimuthu-Muthuvel-2/publication/353122087_Stroke_Prediction/links/61d95d8bda5d105e55278aba/Stroke-Prediction.pdf?_sg%5B0%5D=started_experiment_milestone&_sg%5B1%5D=started_experiment_milestone&origin=journalDetail&_rtd=e30%3D |
| Stroke Article 42 | A Hybrid Deep Learning Approach for Improved Detection and Prediction of Brain Stroke | 10.3390/app15094639 |
| Stroke Article 43 | Prediction of Stroke Disease Using Deep CNN Based Approach | 10.12720/jait.13.6.604-613 |
| Stroke Article 44 | An Interpretable Approach with Explainable AI for Heart Stroke Prediction | 10.3390/diagnostics14020128 |
| Stroke Article 45 | Multi-Layer Perceptron For Diagnosing Stroke With The SMOTE Method In Overcoming Data Imbalances | 10.37058/innovatics.v5i1.6565 |
| Stroke Article 46 | An effective PO-RSNN and FZCIS based diabetes prediction and stroke analysis in the metaverse environment | 10.1038/s41598-025-96541-2 |
| Stroke Article 47 | Stroke Prediction based on Integrated Models of Biological Risk Factors | 10.1145/3644116.3644171 |
| Stroke Article 48 | A Glimpse to the Future: Identifying Stroke Risk Factors Using Data Visualization for Stroke Prediction | 10.56741/esl.v3i01.470 |
| Stroke Article 49 | A hybrid system to predict brain stroke using a combined feature selection and classifier | 10.1016/j.imed.2023.06.002 |
| Stroke Article 50 | Revolutionizing heart attack prognosis: Introducing an innovative regression model for prediction | 10.1016/j.imu.2025.101664 |
| Stroke Article 51 | An Ensemble Machine Learning and Data Mining Approach to Enhance Stroke Prediction | 10.3390/bioengineering11070672 |
| Stroke Article 52 | Intelligent Stroke Disease Prediction Model Using Deep Learning Approaches | 10.1155/2024/4523388 |
| Stroke Article 53 | Combinations of Optimization Method and Balancing Technique in Hypertension Classification with Machine Learning | 10.35882/ijeeemi.v7i2.86 |
| Stroke Article 54 | Stroke Prediction with Enhanced Gradient Boosting Classifier and Strategic Hyperparameter | 10.30812/matrik.v23i2.3555 |
| Stroke Article 55 | Web Design for Stroke Early Detection Using Decision Tree C5.0 | 10.33751/komputasi.v20i2.8265 |
| Stroke Article 56 | A machine learning approach for predicting stroke | 10.53388/MDM202407015 |
| Stroke Article 57 | Explainable and Interpretable Model for the Early Detection of Brain Stroke Using Optimized Boosting Algorithms | 10.3390/diagnostics14222514 |
| Stroke Article 58 | Comparison of Classification and Regression Model Approaches on the Main Causes of Stroke with Symbolic Regression Feyn Qlattice | 10.59247/jahir.v1i2.87 |
| Stroke Article 59 | Improving stroke risk prediction by integrating XGBoost, optimized principal component analysis, and explainable artificial intelligence | 10.1186/s12911-025-02894-z |
| Stroke Article 60 | Predictive modelling and identification of key risk factors for stroke using machine learning | 10.1038/s41598-024-61665-4 |
| Stroke Article 61 | SPE: Ensemble Hybrid Machine Learning Model for Efficient Diagnosis of Brain Stroke towards Clinical Decision Support System (CDSS) | https://ijisae.org/index.php/IJISAE/article/view/2544 |
| Stroke Article 62 | Pre-Diagnosing the Stroke Using Artificial Neural Network | 10.14704/nq.2022.20.10.NQ55093 |
| Stroke Article 63 | Unveiling the potential of machine learning approaches in predicting the emergence of stroke at its onset: a predicting framework | 10.1038/s41598-024-70354-1 |
| Stroke Article 64 | Comparison of Machine Learning and Deep Learning Techniques for Stroke Prediction | 10.29137/umagd.1432162 |
| Stroke Article 65 | BrainOK: Brain Stroke Prediction using Machine Learning | https://www.jetir.org/papers/JETIR2204518.pdf |
| Stroke Article 66 | Enhancing stroke prediction using the waikato environment for knowledge analysis | 10.11591/ijai.v13.i3.pp3010-3017 |
| Stroke Article 67 | Robust classification model for identifying stroke patients utilising a machine learning-based ensemble stacking method | 10.1088/2631-8695/adad3a |
| Stroke Article 68 | Machine Learning Prediction of Brain Stroke at an Early Stage | 10.24996/ijs.2023.64.12.39 |
| Stroke Article 69 | Health Seek: A Deep Learning-Based Intelligent System to Aid Medical Diagnosis | 10.4236/jbise.2022.151007 |
| Stroke Article 70 | Classification of stroke patients using data mining with AdaBoost, Decision Tree and Random Forest models | 10.33096/ilkom.v14i3.1328.218-228 |
| Stroke Article 71 | A new stroke prediction model combined algorithm based on artificial neural networks and logistic regression | 10.1117/12.2656802 |
| Stroke Article 72 | Prediction of Stroke Disease with Demographic and Behavioural Data Using Random Forest Algorithm | 10.3390/analytics2030034 |
| Stroke Article 73 | Enhancing Stroke Prediction Using LightGBM With SMOTE-ENN and Fine-Tuning: A Comprehensive Analysis | 10.7759/s44389-02402268-y |
| Stroke Article 74 | A Robust Methodology for Stroke Disease Prediction using a Hard Voting Classifier | 10.48084/etasr.10292 |
| Stroke Article 75 | An intelligent learning system based on electronic health records for unbiased stroke prediction | 10.1038/s41598-024-73570-x |
| Stroke Article 76 | A brain stroke detection model using soft voting based ensemble machine learning classifier | 10.1016/j.measen.2023.100871 |
| Stroke Article 77 | A Machine Learning Ensemble Classifier for Prediction of Brain Strokes | 10.14569/IJACSA.2022.0131232 |
| Stroke Article 78 | Detecting Stroke in Human Beings Using Machine Learning | 10.54941/ahfe1003460 |
| Stroke Article 79 | A novel K-nearest neighbor classifier for lung cancer disease diagnosis | 10.1007/s00521-024-10235-w |
| Stroke Article 80 | Detecting Brain Stroke Using Enhanced Ensemble Models in Data-Driven Healthcare Systems for the Future Factories | 10.7759/s44389-025-06211-7 |
| Stroke Article 81 | Analyzing the Performance of Stroke Prediction using ML Classification Algorithms | 10.14569/IJACSA.2021.0120662 |
| Stroke Article 82 | Multiple Explainable Approaches to Predict the Risk of Stroke Using Artificial Intelligence | 10.3390/info14080435 |
| Stroke Article 83 | Deep Learning and Machine Learning for Early Detection of Stroke and Haemorrhage. | 10.32604/cmc.2022.024492 |
| Stroke Article 84 | Genetic Folding (GF) Algorithm with Minimal Kernel Operators to Predict Stroke Patients | 10.1080/08839514.2022.2151179 |
| Stroke Article 85 | Process Mining Organization (PMO) Based on Machine Learning Decision Making for Prevention of Chronic Diseases | 10.3390/eng5010015 |
| Stroke Article 86 | Risk factor identification for stroke prognosis using machine-learning algorithms | 10.5455/jjcit.71-1652725746 |
| Stroke Article 87 | Machine Learning Approaches in Medical Data Processing: A Proposal for an Intelligent Stroke Diagnosis System | 10.62520/fujece.1694558 |
| Stroke Article 88 | A Machine Learning Framework for Stroke Prediction: Balancing Precision and Recall in Healthcare Analytics | 10.1109/MeMeA65319.2025.11068036 |
| Stroke Article 89 | Data Management Analysis for Predicting Stroke using RapidMiner | 10.61487/jiste.v2i3.95 |
| Stroke Article 90 | Optimization of Machine Learning Algorithms with Bagging and AdaBoost Methods for Stroke Disease Prediction | https://ami.info.umfcluj.ro/index.php/AMI/article/view/927 |
| Stroke Article 91 | A comparative analysis of machine learning classifiers for stroke prediction: A predictive analytics approach | 10.1016/j.health.2022.100116 |
| Stroke Article 92 | Analysis of Data and Feature Processing on Stroke Prediction using Wide Range Machine Learning Model | 10.15575/join.v9i1.1249 |
| Stroke Article 93 | An Improved Concatenation of Deep Learning Models for Predicting and Interpreting Ischemic Stroke | 10.1109/ACCESS.2024.3386220 |
| Stroke Article 94 | A Predictive Model of Stroke Diseases using Machine Learning Techniques | 10.35940/ijrte.A6900.0511122 |
| Stroke Article 95 | Analysis of Stroke Classification Using Random Forest Method | 10.33096/ilkom.v14i3.1252.186-193 |
| Stroke Article 96 | Effective Stroke Prediction using Machine Learning Algorithms | 10.34104/ajeit.024.026036 |
| Stroke Article 97 | Comparison of Classification Algorithm in Predicting Stroke Disease | 10.34012/jurnalsisteminformasidanilmukomputer.v6i1.2714 |
| Stroke Article 98 | Stroke Prediction | 10.17265/2159-5275/2021.06.004 |
| Stroke Article 99 | Enhanced stroke prediction using stacking methodology (ESPESM) in intelligent sensors for aiding preemptive clinical diagnosis of brain stroke | 10.1016/j.measen.2024.101108 |
| Stroke Article 100 | A novel adaptive weight bi-directional long short-term memory (AWBi-LSTM) classifier model for heart stroke risk level prediction in IoT | 10.7717/peerj-cs.2196 |
| Stroke Article 101 | The CLASSIFICATION OF STROKE PREDICTION USING THE SUPPORT VECTOR MACHINE (SVM) METHOD | 10.35957/jatisi.v11i3.8044 |
| Stroke Article 102 | RDET stacking classifier: a novel machine learning based approach for stroke prediction using imbalance data | 10.7717/peerj-cs.1684 |
| Stroke Article 103 | Stroke prediction based on multifactorial regression models | 10.1117/12.2662578 |
| Stroke Article 104 | Stroke Risk Prediction with Machine Learning Techniques | 10.3390/s22134670 |

List of all the included clinical prediction models articles for the research project using the diabetes dataset.

| Reference Title | Article Title | DOI |
| --- | --- | --- |
| Diabetes Article 1 | Stacking with Recursive Feature Elimination-Isolation Forest for classification of diabetes mellitus | 10.1371/journal.pone.0302595 |
| Diabetes Article 2 | Towards a Stacking Ensemble Model for Predicting Diabetes Mellitus using Combination of Machine Learning Techniques | 10.14569/IJACSA.2023.0141236 |
| Diabetes Article 3 | Modelo predictivo de diabetes utilizando el proceso CRISP-DM para la prevención de la enfermedad implementando Machine Learning | 10.17981/cesta.04.02.2023.02 |
| Diabetes Article 4 | Integrated bagging-RF learning model for diabetes diagnosis in middle-aged and elderly population | 10.7717/peerj-cs.2436 |
| Diabetes Article 5 | Performance Evaluation of Supervised Machine Learning Classifiers for Type 2 Diabetes Mellitus Prediction | 10.34248/bsengineering.1618267 |
| Diabetes Article 6 | Diabetes Risk Prediction Model Based on Improved XGBoost with Genetic Algorithm | 10.1145/3727993.372805 |
| Diabetes Article 7 | Enhancing Healthcare: Machine Learning for Diabetes Prediction and Retinopathy Risk Evaluation | 10.14569/IJACSA.2024.0150703 |
| Diabetes Article 8 | Effective Diabetes Prediction using an IoT-based Integrated Ensemble Machine Learning Framework | 10.48084/etasr.8869 |
| Diabetes Article 9 | Prediction of diabetes using hybrid support vector machines with Artificial Bee Colony | 10.1007/s13410-025-01478-x |
| Diabetes Article 10 | Hi-Le and HiTCLe: Ensemble Learning Approaches for Early Diabetes Detection Using Deep Learning and Explainable Artificial Intelligence | 10.1109/ACCESS.2024.3398198 |
| Diabetes Article 11 | A Machine Learning-Based Prediction Study for Type 2 Diabetic Mellitus | 10.61173/zr192s24 |
| Diabetes Article 12 | Formulation of a Multi-Disease Comorbidity Prediction Framework: A Data-Driven Case Analysis on of Diabetes, Hypertension, and Cardiovascular Risk Trajectories | 10.32996/jcsts.2023.5.3.12 |
| Diabetes Article 13 | Leveraging Data Mining Techniques for Presymptomatic Diabetes Likelihood Prediction | https://share.google/z5yGAkiGRExbJxTzK |
| Diabetes Article 14 | Integrated Ensemble Model for Diabetes Mellitus Detection | 10.14569/IJACSA.2024.0150423 |
| Diabetes Article 15 | Advancing Diabetes Prediction: A Nuanced Six-Class Classification System and Risk Factor Interactions Investigation | 10.2991/978-94-6463-300-9_71 |
| Diabetes Article 16 | Prediction of Type-II Diabetic through Blockchain Technology | 10.17485/IJST/v18i15.3423 |
| Diabetes Article 17 | Predicting Diabetes Mellitus with Machine Learning Techniques | 10.58564/IJSER.4.2.2025.315 |
| Diabetes Article 18 | An effective PO-RSNN and FZCIS based diabetes prediction and stroke analysis in the metaverse environment | 10.1038/s41598-025-96541-2 |
| Diabetes Article 19 | Development of Diabetes Diagnosis Tool Using Machine Learning | 10.30880/eeee.2024.05.01.001 |
| Diabetes Article 20 | A predictive machine learning framework for diabetes | 10.31127/tuje.1434305 |
| Diabetes Article 21 | Data to Diagnosis: Evaluating Machine Learning Algorithms for Predictive Healthcare in Diabetes | 10.21015/vtse.v13i3.2141 |

Articles which have an active web application that is currently publicly available as of 14/01/2026

Stroke Article 38 has publicly available prediction model website: <https://strokedetection.streamlit.app/>

Stroke Article 55 has publicly available prediction model website: <https://rezaummamnor.github.io/StrokePredictionRezaUmmam/>

Medical device patent: <https://worldwide.espacenet.com/patent/search/family/095562296/publication/WO2025097042A1?q=pn%3DWO2025097042A1>
