## Supplementary figures and images for "Evidence of Unreliable Data and Poor Data Provenance in Clinical Prediction Model Research and Clinical Practice"

### Supplementary 3

Diabetes Webpage Screenshots

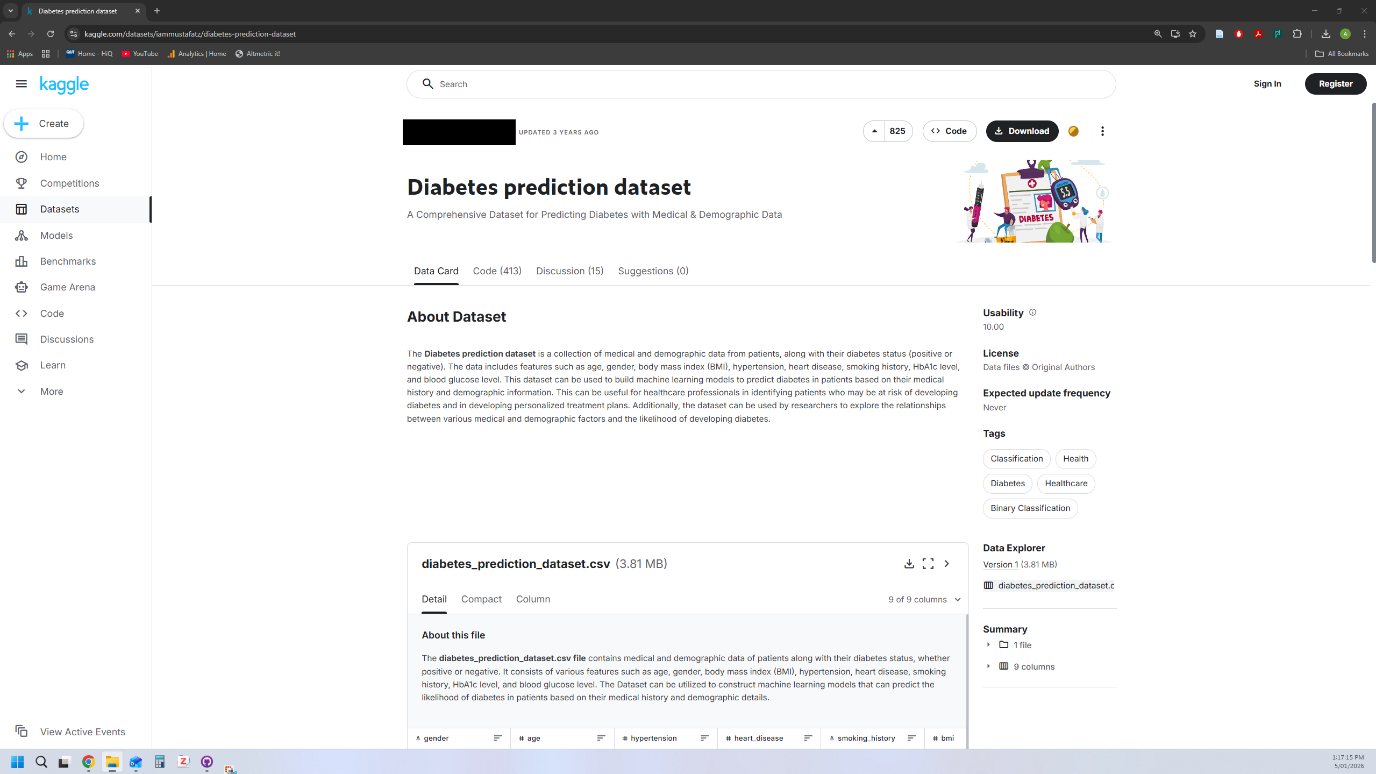

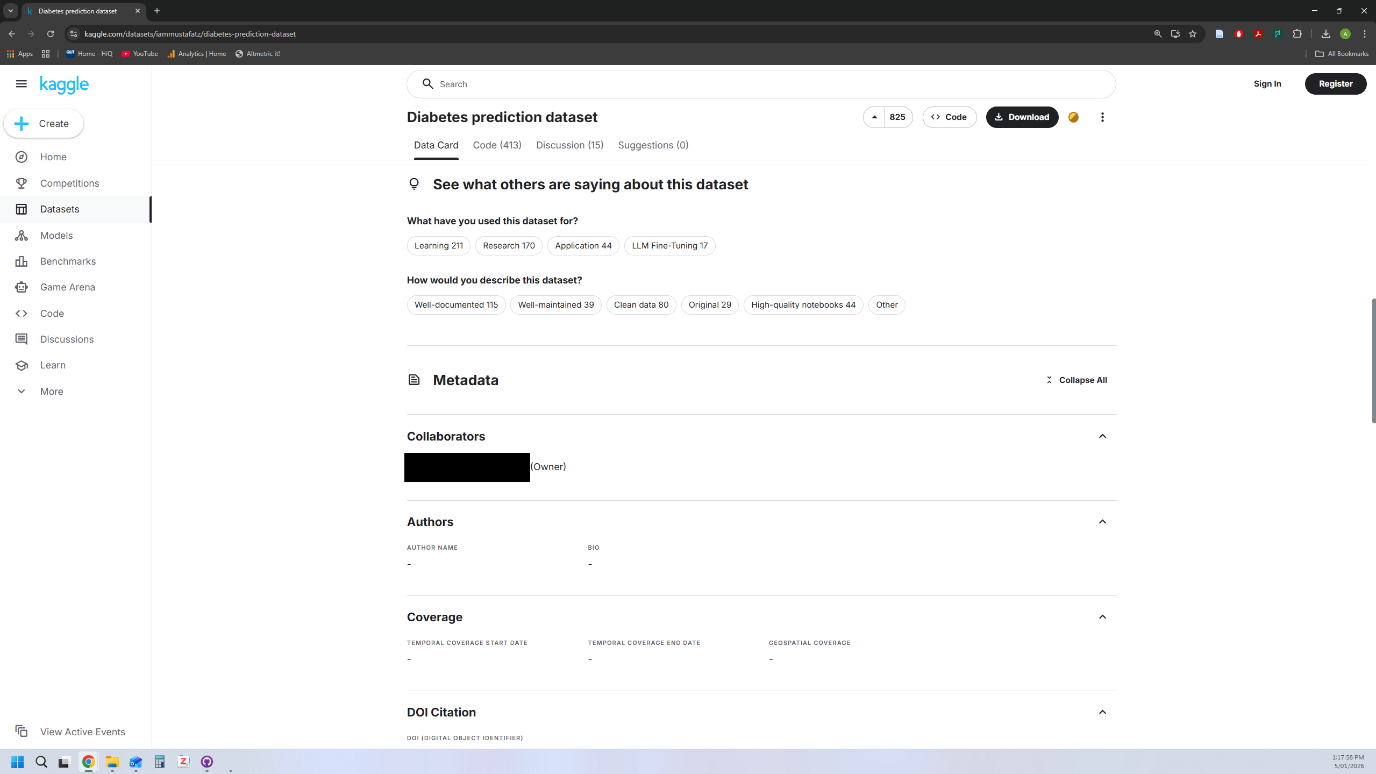

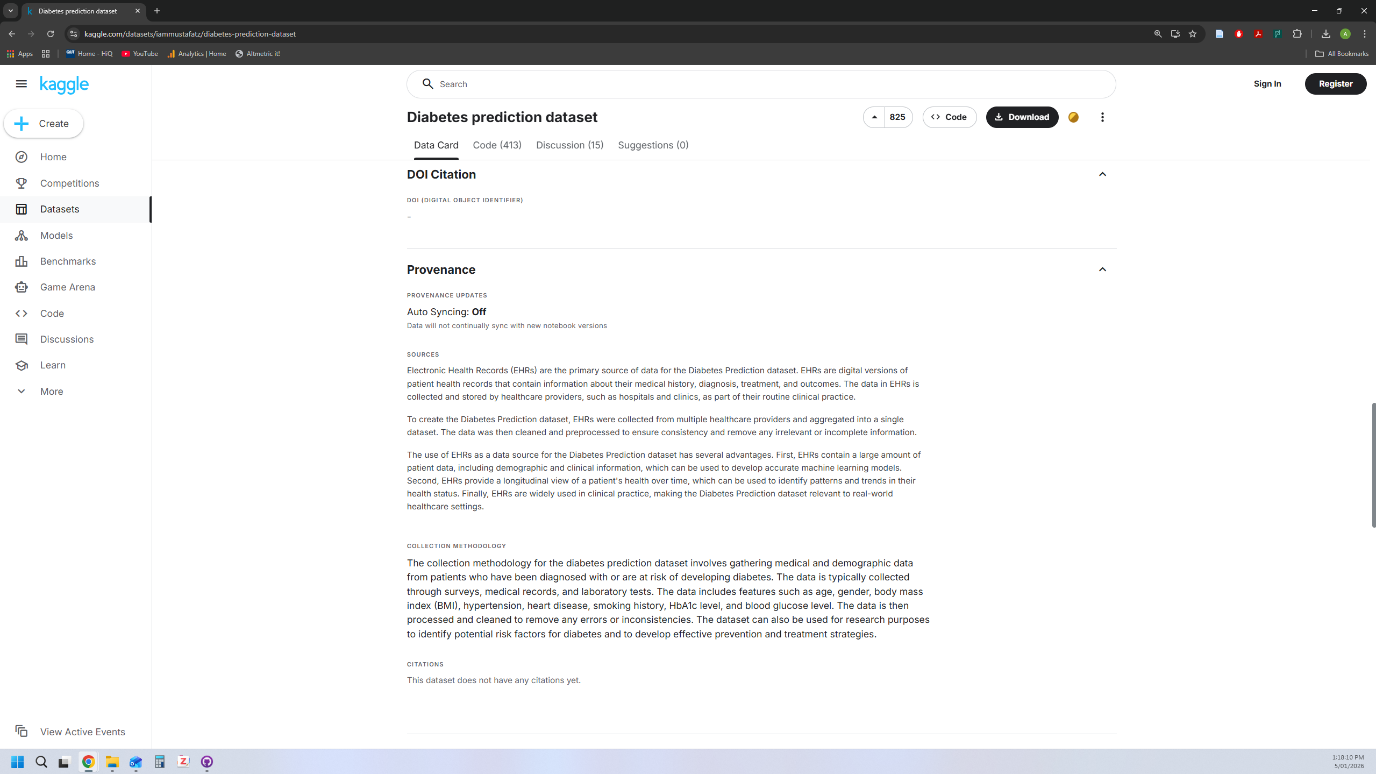

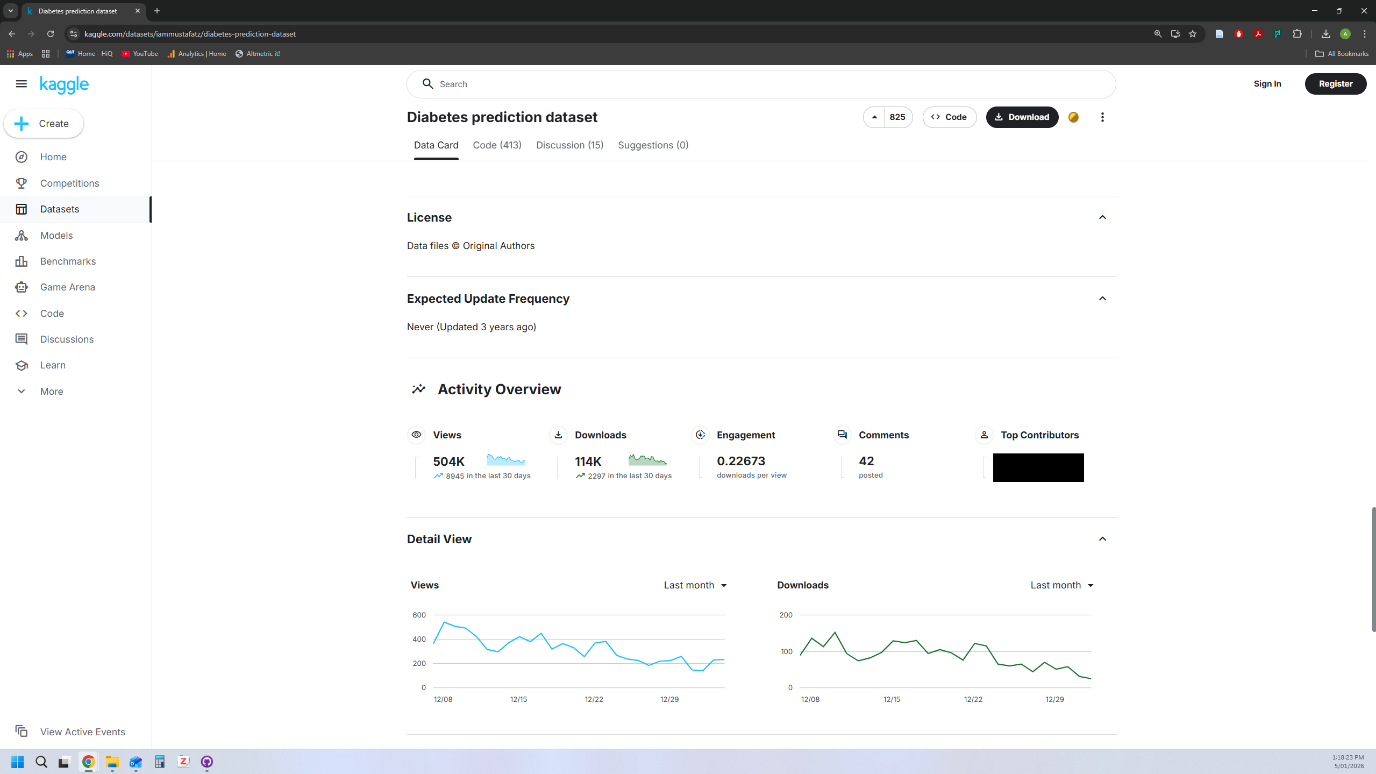
