## Supplementary 1 for "Evidence of Unreliable Data and Poor Data Provenance in Clinical Prediction Model Research and Clinical Practice"

**TRIPOD+AI Checklist items used**

| **TRIPOD+AI Item** | **Description** |
| --- | --- |
| 5a | Describe the sources of data separately for the development and evaluation datasets (e.g., randomised trial, cohort, routine care or registry data), the rationale for using these data, and representativeness of the data |
| 5b | Specify the dates of the collected participant data, including start and end of participant accrual; and, if applicable, end of follow-up |
| 6a | Specify key elements of the study setting (e.g., primary care, secondary care, general population) including the number and location of centres |
| 7 | Describe any data pre-processing and quality checking, including whether this was similar across relevant sociodemographic groups |
| 8a | Clearly define the outcome that is being predicted and the time horizon, including how and when assessed, the rationale for choosing this outcome, and whether the method of outcome assessment is consistent across sociodemographic groups |
| 9a | Describe the choice of initial predictors (e.g., literature, previous models, all available predictors) and any pre-selection of predictors before model building |
| 9b | Clearly define all predictors, including how and when they were measured (and any actions to blind assessment of predictors for the outcome and other predictors) |
| 10 | Explain how the study size was arrived at (separately for development and evaluation), and justify that the study size was sufficient to answer the research question. Include details of any sample size calculation |
| 11 | Describe how missing data were handled. Provide reasons for omitting any data |

**Table 1.** Nine items from the TRIPOD+AI checklist that relate to data provenance and quality which were used to evaluate the dataset information and article adherence (5). The full TRIPOD+AI checklist has 27 items.

**Datasets**

There are two datasets used from <https://www.kaggle.com> for this study.

- The first dataset is for stroke prediction and is publicly available from: <https://www.kaggle.com/datasets/fedesoriano/stroke-prediction-dataset>
- The second dataset is for diabetes prediction and is publicly available from: <https://www.kaggle.com/datasets/iammustafatz/diabetes-prediction-dataset>

**Search terms**

To search for research using either of the datasets we completed searches in Google Scholar. We used information relating to the Kaggle web addresses above to search in Google Scholar.

- The search term for the stroke prediction dataset is: “fedesoriano stroke prediction dataset”
- The search term for the diabetes prediction dataset is: “iammustafatz diabetes prediction dataset”
